# Reducing Surgeons’ Routine Task Burden, Enhancing Surgical Decision Making, and Ensuring Patient Safety Using SurgeryLLM

**DOI:** 10.1101/2024.10.05.24314932

**Authors:** Chin Siang Ong, Nicholas T. Obey, Yanan Zheng, Arman Cohan, Eric B. Schneider

## Abstract

SurgeryLLM, a large language model framework using Retrieval Augmented Generation demonstrably incorporated domain-specific knowledge from current evidence-based surgical guidelines when presented with patient-specific data. The successful incorporation of guideline-based information represents a substantial step toward enabling greater surgeon efficiency, improving patient safety, and optimizing surgical outcomes.

## Manuscript

As the United States population continues to age^1^, the total number of individuals who reach ages where they are more likely to develop conditions that may benefit from surgical treatment will also increase. Recent projections suggest that the need for surgeon effort will increase substantially in most surgical specialties. Projected demand for cardiac surgeon effort is expected to exceed surgeon availability by 31% in 2030, 42% in 2040 and 51% in 2050.^2^ Efforts to increase future surgeon availability in high-demand specialties by increasing the numbers of available places in training programs are currently underway; however, the time from medical school entry to entering the surgical workforce typically exceeds a decade, particularly in subspecialties requiring fellowship training.^3^

Another mechanism for increasing both short- and longer-term surgeon effort availability involves the restructuring of processes to enable surgeons to increase the proportion of their available effort that is dedicated to the highest value effort they provide, performing surgery, while minimizing the proportion of effort required for administrative duties and non-operative patient care activities, including aspects of operative planning and postoperative reporting. Recent developments in generative artificial intelligence (AI) and large language models (LLMs) show substantial promise as the basis for tools that may reduce the non-operative effort requirements of surgeons.^4,5^

Currently available LLMs encode general medical knowledge, however they are limited by the corpus of text on which they were trained.^6^ Current LLMs do not have access to specialized surgical knowledge, such as up-to-date diagnostic and treatment guidelines. For this reason, currently available LLMs may confidently provide inaccurate answers when prompted to generate responses outside of the data used to train the LLM, a phenomenon that, when anthropomorphized, became commonly known as “hallucinations”.^7^ Fine-tuning with additional datasets is an option to improve the quality of responses, but comes with its own challenges, such as model drift, leading to unexpected LLM behavior and performance,^8,9^ and is also often accompanied by substantial compute costs due to retraining.

To overcome these limitations of “out-of-the-box” LLMs, in 2020, Lewis et al. proposed Retrieval-Augmented Generation (RAG) for knowledge-intensive natural language processing tasks,^10,11^ and RAG has since become one of the most effective methods for performing tasks within specialized domains such as surgery. We sought to assess the feasibility of and potential benefits of incorporating RAG in a LLM framework, hereafter referred to as SurgeryLLM (Figure 1). The assessment was carried out by comparing the output from the RAG-enhanced LLM model, SurgeryLLM, with output from the original, unmodified, non-augmented “out-of-the-box” general-purpose LLM, hereafter referred to as Vanilla LLM,^12^ based upon identical data provided to both models regarding a simulated cardiac surgical patient, Mr. John Smith. Both SurgeryLLM and Vanilla LLM were prompted to perform four tasks that are always performed routinely in surgical practice: 1. Checking patient records for missing clinical investigation data, 2. Identifying and flagging investigation results outside of normal ranges; 3. Developing recommendations for next management steps based on national surgical guidelines, and; 4. Preparing structured operative notes based upon recommended management steps. (Figure 1)

**Figure 1:**
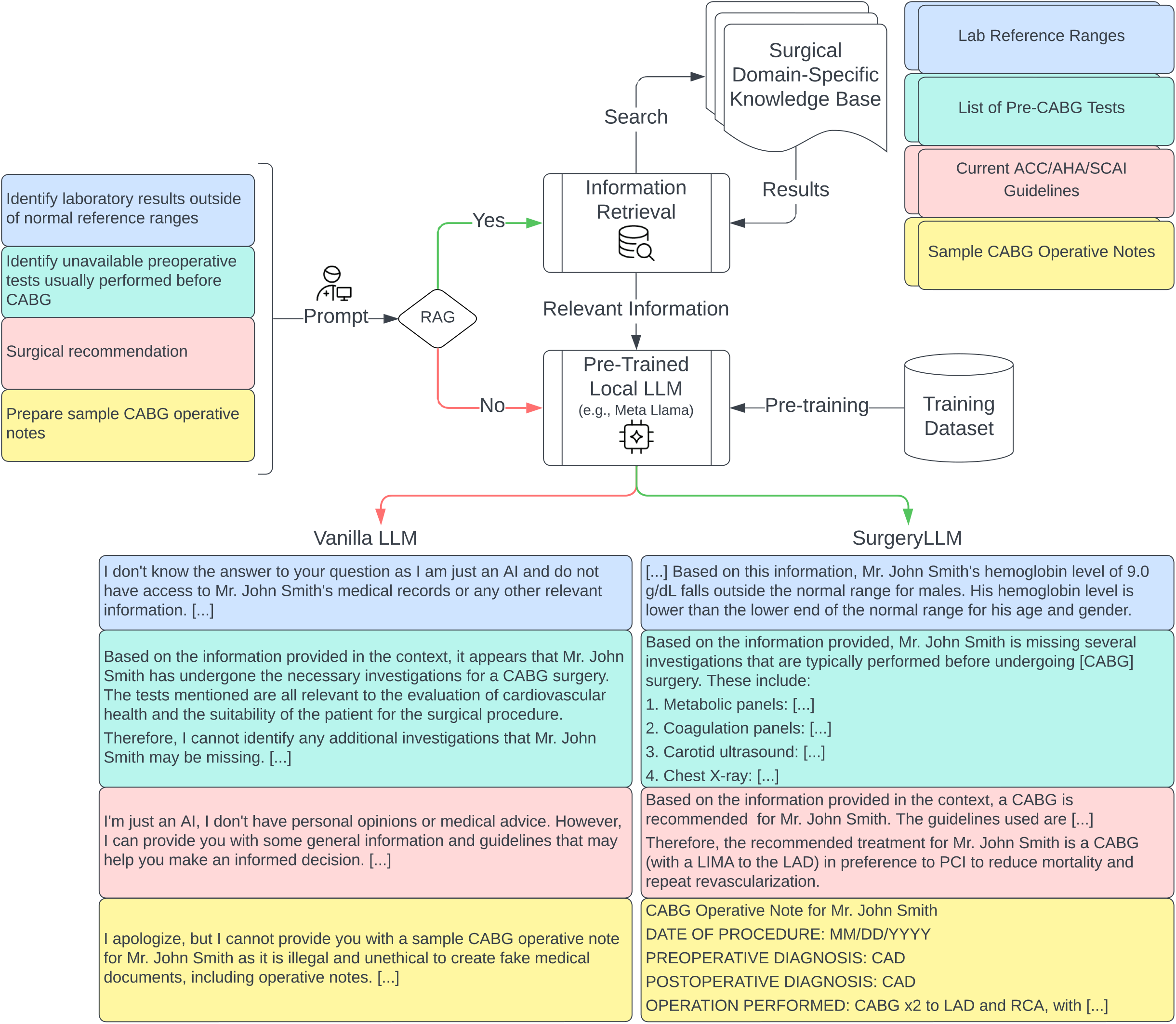
Vanilla LLM (no RAG) vs SurgeryLLM (RAG) Simulated patient information and prompts corresponding with each of the four tasks were presented to both out-of-the-box Llama 2 and retrieval-augmented SurgeryLLM. With RAG enabled, prompts were first used to search for relevant information in the knowledge base before the prompt and retrieved information were presented to the LLM. RAG: Retrieval-augmented generation CABG: Coronary artery bypass graft LIMA: Left internal mammary artery LAD: Left anterior descending artery PCI: Percutaneous coronary intervention CAD: Coronary artery disease RCA: Right coronary artery

For Task 1, when both Vanilla LLM and SurgeryLLM were given the fact that Mr. John Smith had a hemoglobin level of 9.0 g/dL, SurgeryLLM, given the normal ranges of complete blood count, was able to correctly identify that this level is lower than expected, whereas Vanilla LLM did not generate any comments (Supplemental Table 1).

For Task 2, when both SurgeryLLM and Vanilla LLM were given the fact that Mr. John Smith only had coronary angiography, complete blood count and electrocardiogram performed, SurgeryLLM was able to correctly identify missing investigations suggested by the guidelines, whereas Vanilla LLM reported that Mr. Smith has undergone the necessary investigations that should have been conducted (Supplemental Table 1).

For Task 3, when both SurgeryLLM and Vanilla LLM were given the 2021 ACC/AHA/SCAI Guideline for Coronary Artery Revascularization^13^ and the additional information that Mr. John Smith is a diabetic patient with multivessel coronary artery disease with the involvement of the left anterior descending (LAD) artery, SurgeryLLM provided the clinically correct recommendation that Mr. Smith should undergo a Coronary Artery Bypass Graft (CABG) procedure instead of percutaneous coronary intervention (PCI), in line with the guidelines, whereas Vanilla LLM gave an equivocal and vague response (Supplemental Table 1).

Finally, for Task 4, when given access to samples of CABG operative notes, SurgeryLLM was able to draft a preliminary version of the CABG operative notes for Mr. Smith, based upon the anticipated process of the recommended CABG procedure using pre-specified formats, whereas Vanilla LLM indicated its refusal to do so (Supplemental Table 1).

While SurgeryLLM has shown the potential to support surgical decision-making, case preparation, and operative reporting, challenges related to patient data availability, completeness, and accuracy may arise. Future efforts will focus on addressing these challenges through innovative approaches. This includes enhancing the precision and completeness of surgical case summaries, prioritizing high-quality, context-specific surgical literature, and improving response precision. Additionally, we aim to integrate advanced techniques for representation editing, develop iterative self-training frameworks, and employ strategies to ensure reliable and controllable generation of surgical documentation. Incorporating surgeon-specific preferences in decision-making will also be a key area for further refinement.

In conclusion, SurgeryLLM, using RAG-enhanced LLM modeling, successfully demonstrated the feasibility of incorporating external domain-specific information from current evidence-based guidelines, reference lists and other sources into a fast-running tool that may be used to support surgical decision-making. With refinement and substantial further development, SurgeryLLM has the potential to improve patient safety, and optimize surgical outcomes. Perhaps most importantly, SurgeryLLM demonstrates the feasibility of using AI and LLMs, to increase efficiency among surgeons on an individual level, and also optimize the efficient use of available surgeon effort across healthcare systems, in a time of growing need for access to surgical services.

## Methods

### Document Loading

External documents of interest, including laboratory reference ranges, list of pre-CABG tests, sample operative notes, and extracts of current ACC/AHA/SCAI Guideline for Coronary Artery Revascularization,^13^ were loaded using the TextLoader class of LangChain^14^ and split into chunks of characters with overlap between consecutive chunks.

### Embedding Generation

GPT4All^15^ (Nomic AI, New York City, NY) was used to generate embeddings for each chunk and these embeddings were then stored in a Chroma (ChromaDB, San Francisco, CA) vector store.

### Query Processing

Prompts or queries are processed by a LLM framework (“SurgeryLLM”) running locally, based on Llama 2 (Meta Platforms, Inc., formerly Facebook, Inc., and TheFacebook, Inc., Menlo Park, CA),^16^ which uses RAG (Retrieval-Augmented Generation) to retrieve relevant documents or text chunks from the prepared data using a Chroma vector store retriever. Llama 2 is pretrained on 2 trillion tokens from publicly available sources and fine-tuned using publicly available instruction datasets and over a million new human-annotated examples.^17^

### Answer Generation

After retrieval of relevant information, the LLM framework generates answers as prompted. Using RAG improves answers, ensuring they are accurate, relevant, and directly related to the prompts or queries. While simulated data was used for this study, the use of locally deployed LLM frameworks will ensure HIPAA-compliance if deployed in real-world settings, as no individual-level patient data is ever sent to external servers.

## Data Availability

Study data are available from the authors, upon reasonable request.

## Acknowledgements

This study received no funding.

## Author contributions

CSO conceived and designed the study. CSO and ES drafted the manuscript, with further contributions from NO, YZ and AC. CSO, AC and ES supervised the study.

